# Wastewater Surveillance of U.S. Coast Guard Installations and Seagoing Military Vessels to Mitigate the Risk of COVID-19 Outbreaks

**DOI:** 10.1101/2022.02.05.22269021

**Authors:** Gregory J. Hall, Eric J. Page, Min Rhee, Clara Hay, Amelia Krause, Emma Langenbacher, Allison Ruth, Steve Grenier, Alexander P. Duran, Ibrahim Kamara, John K. Iskander, Dana L. Thomas, Edward Bock, Nicholas Porta, Jessica Pharo, Beth A. Osterink, Sharon Zelmanowitz, Corinna M. Fleischmann, Dilhara Liyanage, Joshua P. Gray

## Abstract

Military training centers may be high risk environments for the spread of disease such as COVID-19. Individuals arrive after traveling from many parts of the country, live in communal settings, and undergo high-interaction training. A pilot study of wastewater testing was initiated in February, 2021 to determine its feasibility as a sentinel surveillance tool in the U.S. Coast Guard for SARS-CoV-2. Wastewater was analyzed for the presence of two viral genes, N and E, and quantified relative to levels of a fecal indicator virus, Pepper Mild Mottle Virus (PMMoV). A stability control, Bovine Syncytial Respiratory Virus vaccine, was added to samples to assess sample stability and degradation. Wastewater data was validated by comparison with concomitant screening and surveillance programs that identified asymptomatic individuals infected with SARS-CoV-2 by diagnostic testing at on site medical clinics using PCR. Elevated levels of SARS-CoV-2 in wastewater were frequently associated with diagnosed cases, and in several instances, led to screenings of asymptomatic individuals that identified infected personnel, mitigating the risk of spread of disease. Wastewater screening also successfully indicated the presence of breakthrough cases in vaccinated individuals. A method for assessing blackwater from Coast Guard vessels was also developed, allowing detection of SARS-CoV-2 virus in shipboard populations. In one instance, virus was detected in the blackwater four weeks following the diagnosis of a single person on a Coast Guard cutter. These data show that wastewater testing is an effective tool for measuring the presence and prevalence of SARS-CoV-2 in military populations so that mitigation can occur and suggest other diseases may be assessed similarly. As a result, the Coast Guard has established three laboratories with wastewater testing capability at strategic locations and is actively continuing its wastewater testing program.

## Introduction

Military installations, particularly training commands, face several high-risk factors for the spread of disease. Individuals arrive from around the United States and international locations, having traveled through commercial carriers, and are housed in barracks holding as many as 100 individuals in a single large room. The individuals are then trained under dense conditions in classrooms and shared living facilities, as well as undergoing training that includes strenuous physical exertion in close proximity to each other and the instructors. In many cases, the most intensive mitigation measures of viral transmission risks such as remote work are not possible without degradation of the training that could affect military readiness. Furthermore, individuals in training are typically young adults who are more likely to have asymptomatic infections (Yonker et al., 2020).

To mitigate the risk of infections, screening measures were implemented prior to and after the arrival of trainees. Restriction of movement (ROM) periods which minimize social contact between individuals are used in congregate settings upon the arrival of personnel at commands together with diagnostic tests before and after arrival at fixed intervals. Infected individuals are then isolated and those in close contact with cases are quarantined to ensure further spread does not occur. Similarly, military vessels feature a crew living in close quarters with one another for extended periods of time punctuated by “port calls” in which individuals may be exposed to communities at locations other than their home port. Outbreaks can quickly erode the ability of a training command to complete its mission; individuals in isolation or quarantine receive degraded training and education and managing the ill could overwhelm the unit’s resources. Therefore, early detection is crucial to prevent outbreaks becoming so large that they severely curtail the mission completion of a base or vessel.

As a larger percentage of the population become vaccinated, diagnostic testing of persons will decline leaving decision-makers with less information about the prevalence of the virus in their communities. As has been noted, some vaccinated people can still shed the virus, particularly variants that evade immunity such as Delta and Omicron, and the virus burden in the community is an important driver for infection risk mitigation policy (Bergwerk et al., 2021; Brown et al., 2021; Paul et al., 2021; Strafella et al., 2021).

Wastewater surveillance is an important tool used for assessing viral load in a population, and can be used to augment other surveillance methods to help close this information gap (Larsen & Wigginton, 2020). Indeed, SARS-CoV-2 is shed in fecal matter due to infection within the small intestine providing a fecal biomarker for infection (Bivins et al., 2020). Many colleges and universities have successfully adopted dormitory-focused wastewater testing programs that screen for the presence of SARs-CoV-2 (Harris-Lovett et al., 2021). As seen with the University of Arizona, as little as a single case can be detected in wastewater and this information can be used to deploy testing to identify infected individuals (Betancourt et al., 2021).

In this paper, we report the implementation of wastewater surveillance for SARS-CoV-2 at three U.S. Coast Guard training installations. We also discuss successes and challenges with wastewater surveillance on military seagoing vessels, specifically Coast Guard cutters.

## Materials and Methods

### Wastewater Sample Processing and RNA Extraction at Coast Guard Installations

Three Coast Guard installations within the continental United States were chosen for wastewater testing, the names of which are redacted and hereafter referred to as Training Centers A, B, and C. Training Center A has a single large barracks housing over 1000 trainees. Most trainees arrive in very large cohorts and remain at the center from August through May, except for shared vacation periods. Training Centers B and C have smaller barracks capable of housing 50-200 individuals, typically. Trainees arrive sporadically throughout the year and stay for varied lengths of time for various training courses. In all three cases, trainees share living spaces, dining facilities, and are frequently associated with one another in close contact.

Wastewater access points were identified downstream of the dormitories/barracks at each base within the United States. To sample the wastewater, a tampon (OB Ultra Absorbency, Edgewell Personal Care, Shelton, CT) was connected by a resilient line attached to a carabiner and placed in the wastewater stream as previously described for 24-96 hours (Corchis-Scott et al., 2021). After retrieval, samples were placed in sealed bags and spiked with a storage control if samples were to be stored for more than one day, Bovine Syncytial Respiratory Virus (BRSV) vaccine (Inforce 3, Zoetis Inc., Kalamazoo, MI). Samples were typically processed within 1-4 days of acquisition. To extract the sample, a corner of the plastic bag was cut, the liquid was extracted into a 50 mL conical centrifuge tube, and the volume was adjusted to ∼48 mL volume with distilled water. The sample was then concentrated using Nanotrap Magnetic Virus Particles (Ceres Nano, Manassas, VA) according to the “Rapid SARS-CoV-2 Viral Isolation from Wastewater” Application Note (Ceres Nano, 2020). RNA was subsequently extracted using the QIAmp Viral RNA Mini Kit (Qiagen, Germantown, MD) according to the manufacturer’s protocol.

### Wastewater Sample Processing and RNA Extraction from Seagoing Vessels

Wastewater streams on seagoing vessels are separated into blackwater (containing fecal and urinary waste) and gray water (containing wastewater from other sources, such as sinks, showers, and kitchen water sources.) Blackwater was sampled from the storage tank or while docked and pumping waste ashore. Blackwater contains significantly more fecal matter than samples acquired from dormitories and cannot be directly extracted using the method as described above. To assess these samples, 5 mL of blackwater was diluted with distilled water to a volume of ∼48 mL, spiked with BRSV process control, and purified using Nanotrap Magnetic Virus Particles as described above.

### Assessment of Viral Genes by Polymerase Chain Reaction

RNA was analyzed by real-time quantitative reverse-transcription polymerase chain reaction. Viral gene N was assessed using CDC-designed primers (N1) (Centers for Disease Control and Prevention, 2020) and the viral E gene was assessed using the primer designed by the Germany Centre for Infection Research (Corman et al., 2020). Pepper Mild Mottle Virus (PMMoV) primer and probe sequences were designed by Eiji Haramoto et al. of the University of Yamanashi, Kofu, Yamanshi, Japan (Haramoto et al., 2013). Bovine Syncytial Respiratory Virus (BRSV) primer and probe sequences were designed by the Veterinary and Agrochemical Research Center, Uccle, Belgium (Boxus, Letellier, & Kerkhofs, 2005). Primers and probes were purchased from Integrated DNA Sciences Inc. (Coralville, IA). Viral genes N1 and E were labeled with the fluorophore FAM, whereas PMMoV and BRSV were labeled with HEX to allow duplex PCR reactions. Primer sequences are shown in Table 1.

**Table 1.**
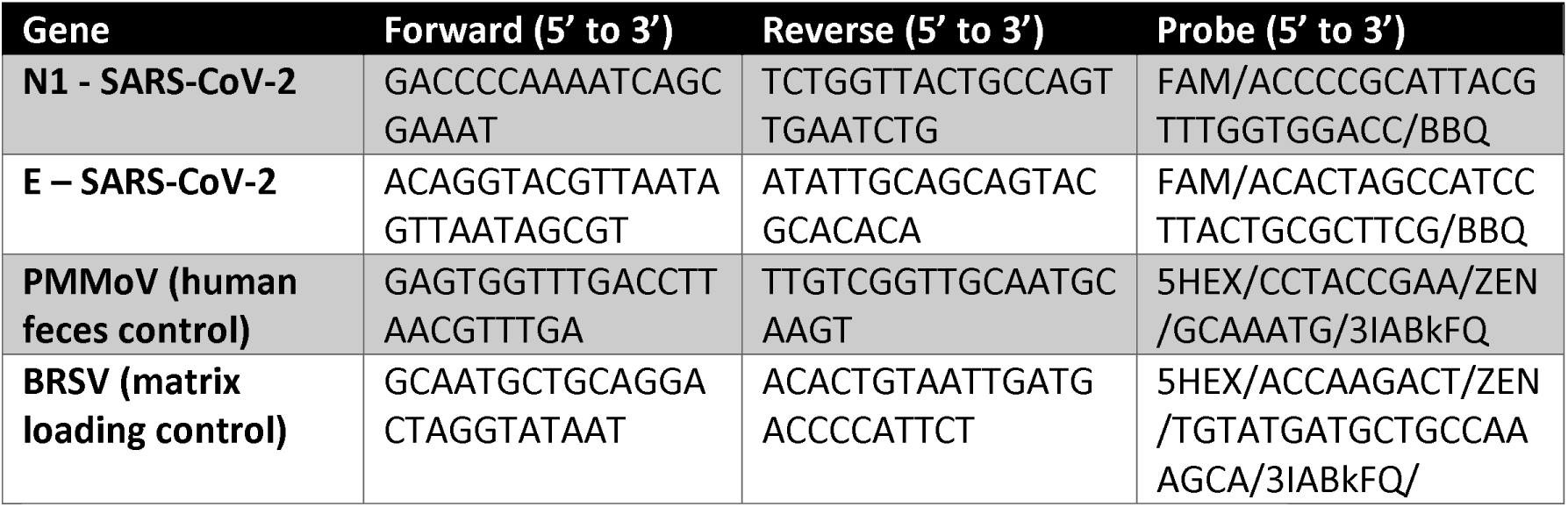
Primer and probe sequences used in quantitative real-time RT-PCR.

One step RT-RT-PCR was performed using TaqPath 1-Step RT-qPCR Master Mix (ThermoFisher Scientific, Waltham, MA) according to manufacturer’s protocol. N1 was multiplexed with BRSV using the following conditions in a CFX-96 real-time PCR thermocycler (Biorad, Hercules, CA): 50C for 10 min, 95 C for 10 min, and 45 cycles of (95 C for 10 s, 60C for 30 s). E was multiplexed with PMMoV using the following conditions: 50C 10 min, 95C 10 min, and 44 cycles of (95C 15 s, 58C, 30 s). C_q_ values were determined by setting the threshold value to 200 (arbitrary fluorescence units).

Amplification of the BRSV process control was used to validate sample stability if stored greater than 24 hours and the absence of inhibitors of PCR in the purified RNA. Samples that did not amplify BRSV were considered failed runs. Positive controls for N1 and E were obtained from Integrated DNA Technologies and were used during each run. The expression of viral genes N and E was compared against expression of PMMoV using the delta-delta Ct method with the assumption of 100% efficiency of PCR amplification (Livak & Schmittgen, 2001). The level of N or E in each sample was quantified relative to PMMoV to estimate the level of virus versus fecal matter in each sample and eliminate confounding factors such as dilution by extraneous sources of wastewater in the samples by gray water or stormwater runoff.

From March through December 2021, the number of each PCR-diagnosed COVID-19 case at each location (barracks and vessel) was compared against the viral burden in the wastewater. Individuals were considered positive for two weeks post-diagnosis if they remained in a sampled population. No identifying information was provided to the authors for any of diagnosed individuals and no studies on humans were performed in this work. Exact dates of testing are redacted to protect the privacy of the individuals, although the time frames are included to allow correlation with national trends in COVID-19 diagnosis at those times, such as the rise of the delta variant.

### Ethical Considerations

The Chair of the Institutional Review Board of the United States Coast Guard Academy, Dr. Leonard M. Giambra, concluded on August 19, 2020 that this work is public health surveillance in accordance with 45 CFR part 46, and therefore is not human research subject to review by an IRB.

## Results and Discussion

Results and discussion are presented for three test sites: Training Centers A, B, and C. Details of these sites are provided in the methods section. Four U.S. Coast Guard cutters were also tested. Names of training centers and cutters are withheld for national security reasons.

### Training Center A

At this training center, a single large barracks houses over 1000 individuals in one, two or three person rooms. Trainees have shared living spaces, restrooms, and a communal dining hall. Wastewater sampling was performed at two wastewater effluent points, each from approximately half of the barracks. Because individuals frequently move between the annexes for meals and restroom use, both effluent points were treated as providing wastewater from the entire building. Individuals diagnosed with COVID-19 were immediately moved to a separate location outside of the barracks and therefore did not contribute to the effluent post-diagnosis. Wastewater sampling was performed from March through December, 2021 and compared with diagnosed cases of COVID-19 in trainees housed in the barracks (Figure 1). Three distinct events occurred during this time period.

**Figure 2.**
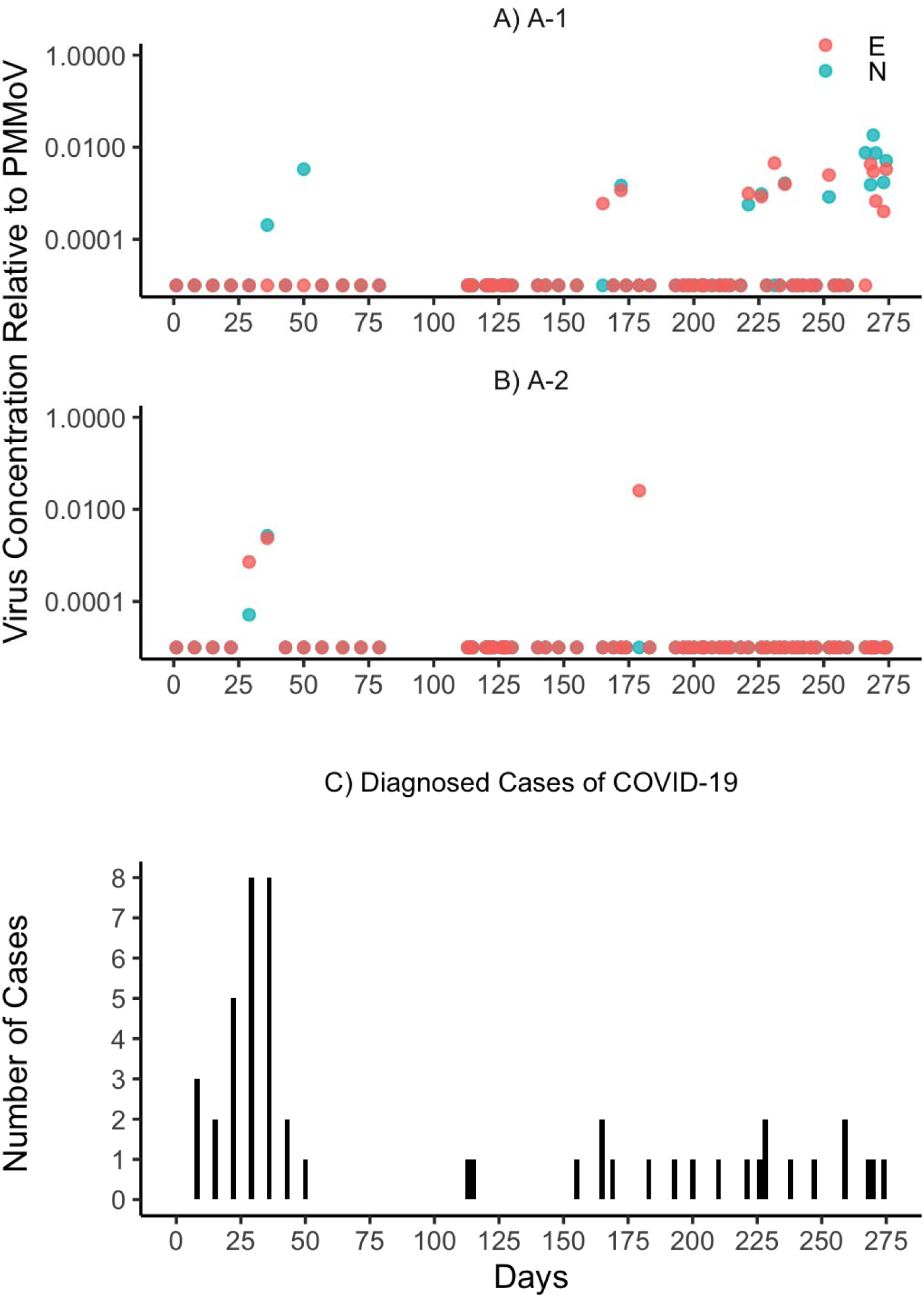
SARS-CoV-2 levels relative to PMMoV at Training Center A. Red (E gene) and blue (N gene) symbols show the level of viral genes relative to PMMoV fecal indicator virus. Samples were taken from March through December for a total of 274 days. Exact sample dates are redacted to protect the privacy of the individuals. Two wastewater sampling sites were measured from the same building. Clinically diagnosed cases are shown in Panel C.

From days 1 through 53, 26 individuals were diagnosed with COVID-19 through diagnostic or surveillance testing, or through contact tracing (Table 2). Wastewater from 24-hour samples analyzed on days 29, 36, and 50 harbored genome fragments for SARS-CoV-2.

**Table 2.** Correlation between diagnosed cases and levels of SARS-CoV-2 genes N and E relative to PMMoV in wastewater effluent from two sampling points in Barracks A. “--” indicates viral genes not detected. Viral load is shown relative to the expression of the fecal indicator PMMoV using the delta-delta Ct method. During this pilot phase, passive samplers were deployed for 24 hours prior to the testing date. Clinically diagnosed individuals were immediately moved to a separate barracks facility and did not contribute to wastewater thereafter.

| Date of wastewater sample | Diagnosed Cases by date | Sampling Point 1 |  | Sampling Point 2 |  |
| --- | --- | --- | --- | --- | --- |
|  |  | N | E | N | E |
| Day 2 | N.A. | -- | -- | -- | -- |
| Day 9 | 3 – day 7<br>Total 3 | -- | -- | -- | -- |
| Day 16 | 1 – day 11<br>1 – day 14<br>Total 2 | -- | -- | -- | -- |
| Day 22 | 4 – day 17<br>1 – day 21<br>Total 5 | -- | -- | -- | -- |
| Day 29 | 2 – day 23<br>2 – day 25<br>2 – day 27<br>4 – day 28<br>Total 10 | -- | -- | 0.0000513 | 0.00072 |
| Day 36 | 2 – day 31<br>4 – day 35<br>1 – day 36<br>Total 7 | 0.000204 | -- | 0.00267 | 0.00236 |
| Day 43 | 2 – day 39<br>Total 2 | -- | -- | -- | -- |
| Day 50 | 1 – day 49<br>Total 1 | 0.003331 | -- | -- | -- |

Because individuals were removed from the population immediately following diagnosis, this may have impacted detection of viral genes from their feces if the test was performed many days later. Indeed, the number of cases and their proximity to the sampling date were hypothesized to play a role in the detection of viral genes. For example, no viral genes were detected on day 22; four cases had been identified five days prior, but only one case one day prior. In contrast, a robust viral detection occurred on day 36; four cases were identified on day 35 and one case on day 36. By mid-April, nearly 95% of the trainees had received their first dose of Pfizer COVID-19 vaccine, and by mid-May, nearly 95% of the individuals at Training Base A were fully vaccinated. Weekly tests continued and no further cases were diagnosed until the end of June.

We concluded that more frequent testing would be performed upon the return of a new cohort of trainees and that placement of the passive samplers in wastewater should occur for the entire interval between sampling days rather than only for 24 hours. Logistically, placing the samplers upon retrieval of the previous sampler was convenient and provided a more longitudinal assessment of wastewater than a 24-hour sample. In addition, because the sampler was already placed, it could be harvested more frequently if needed, such as if a large number of cases were diagnosed or elevated levels of SARS-CoV-2 gene fragments were detected. Thus, the time period each sampler was deployed varied based on when it was retrieved, but the placement typically occurred for 24-96 hours.

A new cohort of trainees from throughout the United States arrived on day 112. The arrival of new cohorts of trainees was considered the most risky time frame, and therefore samples were taken on days 113, 114, and 115. Although two cases of COVID-19 were clinically diagnosed on day 112 and day 115, SARS-CoV-2 was not detected in the wastewater. Wastewater sampling was adjusted from one 24-hour taken once a week to two 72-96 hour samples taken approximately twice per week thereafter.

From July through mid-August, no cases of COVID-19 were clinically diagnosed and no virus was detected in the wastewater. A new cohort of trainees arrived in mid-August, and from that time until present, individuals were tested weekly with antigen tests for COVID-19. The vaccination rate in the cohort was greater than 95%. From mid-August through mid-October, 9 cases of COVID-19 were clinically diagnosed. SARS-CoV-2 gene fragments were only detected on four occasions: days 165, 172, 179, and 221. In each of these instances, a COVID-19 case was clinically diagnosed within four days before or after the detection.

Two interesting outcomes occurred from September-December. The highest level of SARS-CoV-2 gene relative to PMMoV in the wastewater detected to that point occurred on day 179 (a ratio of 0.0254), but no virus was detected in the wastewater on the subsequent measurement, day 183. A clinical diagnosis was made on day 183 for a trainee. It was determined that the trainee had spent a few days away from the barracks, explaining the loss of signal on day 183.

The second event occurred during a second post-holiday time frame (Table 3). Daily wastewater tests were performed in conjunction with once-weekly testing of the entire population. Wastewater levels reached a very high level (a SARS-CoV-2 N gene ratio to PMMoV of 0.0183) on day 269. Three cases were diagnosed the first week and one the second week, with the individuals moved to isolation in a separate building. We concluded that individual cases, largely in vaccinated individuals, shed amounts of virus that were at or near the lower limit of detection for our assay in this population of more than 1000 individuals at this particular location. Additionally, the sustained signal post-holiday suggests that additional individuals are shedding despite testing negative on two COVID-19 antigen tests.

**Table 3.** Correlation between diagnosed cases and levels of SARS-CoV-2 genes N and E relative to PMMoV in wastewater effluent from two sampling points in Barracks A following a holiday weekend. “--” indicates viral genes not detected. Viral load is shown relative to the expression of the fecal indicator PMMoV using the delta-delta Ct method. During this phase, passive samplers were deployed for 24 hours or longer, from the time of removal of the last passive sampler. Clinically diagnosed individuals were immediately moved to a separate barracks facility and did not contribute to wastewater thereafter.

| Day of wastewater sample | Day of Diagnosed Cases | Sampling Point 1 |  | Sampling Point 2 |  |
| --- | --- | --- | --- | --- | --- |
|  |  | N | E | N | E |
| 266 | N.A. | 0.00760 | -- | -- | -- |
| 268 | 1 – day 268 | 0.00153 | 0.00428 | -- | -- |
| 269 | 1 – day 269 | 0.01833 | 0.00298 | -- | -- |
| 270 | 1 – day 270 | 0.00749 | 0.00068 | -- | -- |
| 273 | N.A. | 0.00172 | 0.0004 | -- | -- |
| 274 | 1 – day 274 | 0.00508 | 0.00040 | -- | -- |
| 275 | N.A. | -- | 0.00059 | -- | -- |
| 276 | N.A. | -- | -- | -- | -- |
| 277 | N.A. | 0.00081 | 0.00057 | -- | -- |
| 280 | N.A. | 0.00107 | -- | -- | -- |

### Training Center B

At Training Center B, individuals live in a dormitory arrangement with two occupants per room, with approximately 100 individuals per building. Wastewater testing was initiated after the establishment of a laboratory capability in June. Each barracks at this location has two wastewater effluent sampling points. Samples were taken once weekly beginning day 4, and twice weekly beginning day 18. By mid-July, the highly transmissible Delta variant became the predominant strain of COVID-19 circulating nationally (Grannis et al., 2021). Unlike the individuals at Training Center A who were over 95% vaccinated, individuals at Training Center B were 58.5% fully vaccinated by day 1 and 79.5% fully vaccinated by day 117. They also arrived and departed for training as staggered cohorts for multiple weeks, increasing the risk of individuals from throughout the United States bringing infections to the base.

Overall, SARS-CoV-2 was detected more frequently and in greater amounts than at Training Center A (Figure 2). Patterns of wastewater detection showed multi-testing day levels that were consistent with arrival and departure of cohorts of trainees. Two of these cases will be highlighted in this discussion. Medical posture at Training Center B allowed for testing of unvaccinated individuals, but did not permit testing of vaccinated individuals except when they were identified as close contacts with a clinically diagnosed individual.

**Figure 2.**
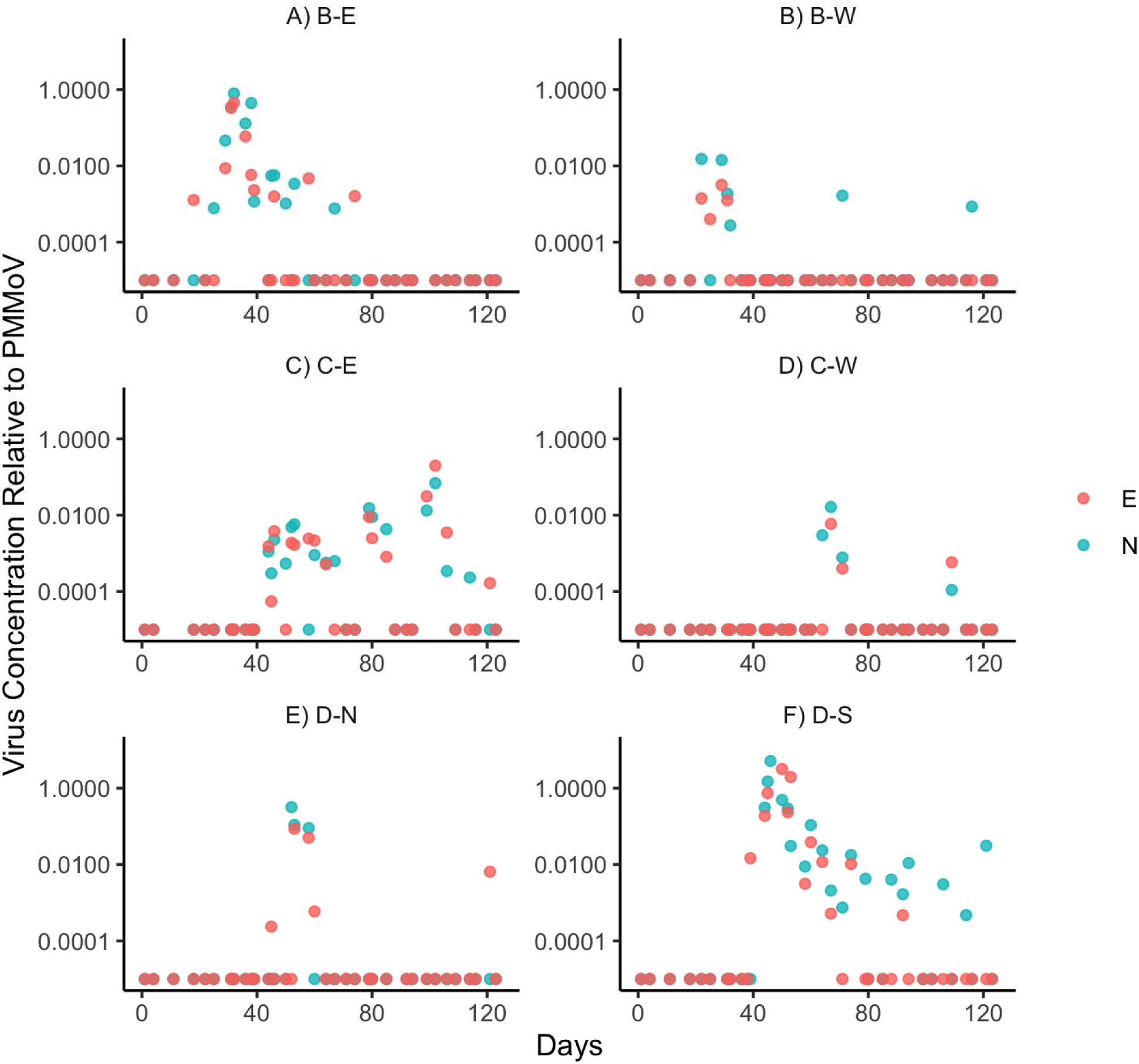
SARS-CoV-2 levels relative to PMMoV at Training Center B. Blue and red symbols show the level of viral genes N and E relative to PMMoV fecal indicator virus. Samples were taken from over a 123-day period from June through October from three buildings, B, C, and D, and two locations, East and West (E and W) or North and South (N and S).

In the first scenario, SARS-CoV-2 genetic fragments were first detected on day 18 in Barracks B-East, with the first surge in viral levels occurring from days 25-39. This spurred testing of all unvaccinated individuals (12 out of 150) in that barracks, but no cases were detected. Wastewater testing was accelerated in response to elevated levels, with wastewater tests performed on July days 31, 32, 36, 38, and 39, yielding some of the highest levels of SARS-CoV-2 ever detected in the study. The individuals living in the barracks finished their training and left on day 38, corresponding with a large drop in detection of viral genes in the wastewater. Vaccinated individuals living in that barracks never became symptomatic and therefore were therefore not clinically tested. It is possible that SARS-CoV-2 was shed by one or more persons experiencing asymptomatic breakthrough infection.

Barracks D-South experienced a rise in SARS-CoV-2 levels beginning on day 39 and ending on day 79; Barracks C North experienced SARS-CoV-2 detection in wastewater from days 52-60. Clinical diagnosed cases of SARS-CoV-2 were made in Barracks D South on day 50, day 52 (2 cases), and day 53 (2 cases). Two clinical diagnoses were made for individuals in Barracks D North on day 53. SARS-CoV-2 remained detectable in wastewater through day 79 in Barracks D South, but no further cases were clinically diagnosed.

Additional surges in SARS-CoV-2 in the wastewater occurred throughout the summer and fall in different barracks locations. Vaccinated individuals residing in a barracks with SARs-CoV-2 in the wastewater were rarely tested. Therefore, vaccinated individuals may have been responsible for prolonged shedding of SARS-CoV-2 observed at these locations, as infected individuals were not identified and removed from a population. Nonetheless, SARS-CoV-2 viral detection was robust and consistent.

### Training Center C

Training Center C is a training center much like Training Center B in that several courses are offered with overlapping schedules with different populations of individuals. Testing began in September and was conducted twice per week with samples being analyzed at the laboratory at Training Center A (Figure 3). Trainees were 73.2% fully vaccinated on day 1 and 83.6% fully vaccinated on day 39. Barracks I houses all students in isolation following a positive clinical diagnosis for COVID-19; that barracks has consistently shown SARS-CoV-2 genetic fragments in the wastewater. The other four barracks each have shown periods of SARS-CoV-2 in the wastewater, and follow-on clinical testing has at times included only unvaccinated individuals. Clinical case numbers by date and barracks were not available at the time of this publication. Barracks H’s wastewater sampling location includes wastewater from Barracks I; a sampling location serving only Barracks H was not identified. Wastewater levels of SARS-CoV-2 relative to PMMoV were similar in Barracks H and I.

**Figure 3.**
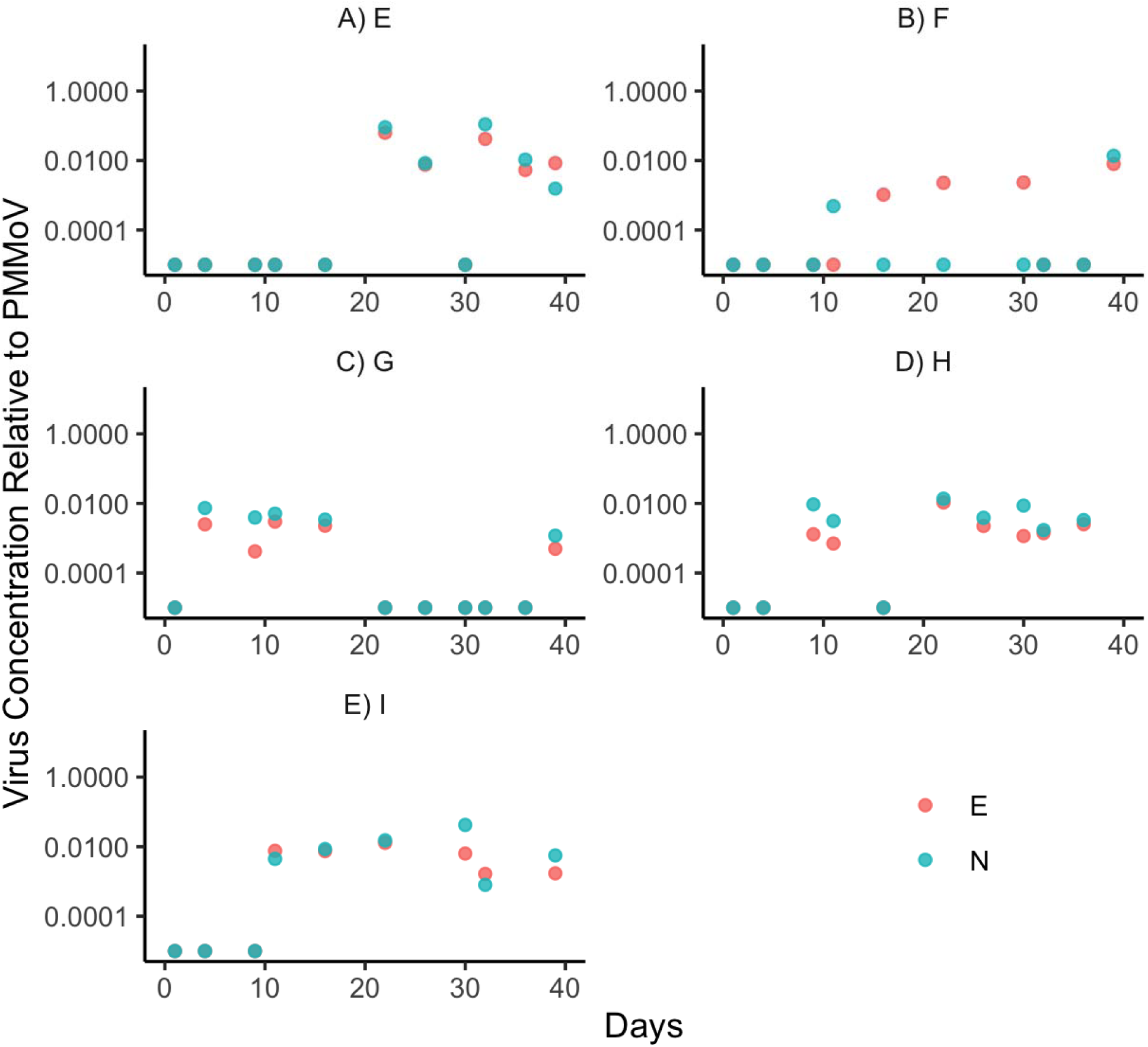
SARS-CoV-2 levels relative to PMMoV at Training Center C. Blue and red symbols show the level of viral genes N and E relative to PMMoV fecal indicator virus. Samples were taken over a 39-day period from September through October.

### Coast Guard Cutters

Sampling wastewater from cutters and other seagoing vessels is more difficult. Although sampling can be done while a vessel is underway, the sample must be stored until the vessel returns to port, limiting its value. Accessing blackwater on the cutters required entering the lowest deck of the ships and opening access points that were sometimes under pressure. Modifications were made to four Coast Guard Cutters to allow for sampling while at dock: a special valve was added that allowed sampling of blackwater as it was pumped from the bilge to dockside septic containers. Multiple samples may need to be taken as the blackwater is not well-mixed. Because it accumulates over various lengths of time, the sample may represent many days’ worth of sample rather than a homogenous mixture from a defined period. This can be addressed by taking multiple samples and homogenizing them at the laboratory.

Despite these challenges, SARS-CoV-2 was detected on two separate occasions. Blackwater is quite concentrated and allows for much greater sensitivity; levels of PMMoV detected were significantly higher than those in barracks. The first was from a cutter for which a known diagnosed case existed on board. In this scenario, an individual was diagnosed using an antigen test and isolated onboard while the cutter continued operations. During the subsequent four weeks prior to arrival at the cutter’s U.S. homeport, the blackwater was dumped at sea on multiple occasions in accordance with standard practices. Upon return to port, four samples were taken while emptying the blackwater storage tank. One of the four samples had genetic fragments for SARS-CoV-2: a value of 2.64E-5 for N1 and 1.96E-5 for E. The results are confounded by the long interval between diagnosis and testing and the subsequent dumping of blackwater several times before the sample was taken. Despite these confounding variables, the outcome shows that SARS-CoV-2 can be detected in blackwater.

SARS-CoV-2 was detected on a separate occasion from a second cutter. The levels detected were 1.21E-4 for N1 and 3.34E-4 for E gene on in June. A subsequent test nearly two weeks later yielded a positive result only for N1, with a value of 2.42E-4. Wastewater triggered a focused diagnostic test of the crew which yielded no positives, but did provide reassurance to begin the subsequent deployment and continue the mission. The vessel had been visited by contractors and guests for a ceremony within the previous week, suggesting a non-crew member was carrying the virus or that an asymptomatic vaccinated person was shedding virus.

## Conclusions and Limitations

This is a report of operational science where we employed the best practices and innovations at the time to provide the leadership of the Coast Guard with timely, scientifically valid advice during the midst of a pandemic. Over the course of this study, wastewater sampling techniques were adjusted to be more frequent and to sample wastewater for longer periods of time than 24 hours. Absent a large outbreak this seems to be a best-practice for communities this size. Evidence from Training Center A indicated that cases had a greater likelihood of not being detected in wastewater if the patients were isolated more than 48 hours prior to a 24 hour assessment, presumably because wastewater would have flushed past the sampling site before placement of the passive sampler. Longer sampling times punctuated by twice or thrice-weekly tests may reduce the risk of false negatives in wastewater.

The passive sampling methodology, the use of a commercial-grade tampon, was chosen for its simple design and inexpensive cost, and because it required minimal expertise to implement (Bivins et al.,2021; Corchis-Scott et al., 2021). Sampling consists of placement of the tampon in the wastewater stream for a period of time, in our case, 24-96 hours. This method detected fecal indicator virus PMMoV 100% of the time. Other institutions have similarly adopted tampons as a passive sampler, including the University of Connecticut from which our method was developed and the University of Arizona (Bivins et al., 2021).

The sensitivity of the method was inversely correlated to the number of individuals living in a barracks. Training Center A, which has a single barracks that houses over 1000 individuals, had a strongest-ever-detected SARS-CoV-2/PMMoV ratio of 0.0254. Training Center B which has dormitories that house 100-150 individuals had the strongest signal of 11.6, nearly three orders of magnitude higher. This finding is also likely to be impacted by the number and strength of infections and the high vaccination rate of the individuals at Training Center A. Due to the dilution of the wastewater by non-infected individuals, the ability to detect single infected individuals is reduced if more individuals live in the same barracks and contribute to single waste stream. Indeed, from April through November, SARS-CoV-2 was detected on eight occasions at Training Center A, the largest barracks in the Coast Guard, despite 21 cases occurring during that timeframe. SARS-CoV-2 viral load, when detected, was much higher at the other bases with smaller dormitories typically housing 150 trainees. These data suggest that a correlation between signal strength and SARS-CoV-2/PMMoV ratio per person may be different at each location. Further confounding variables are that the amount of SARS-CoV-2 shedding by individuals is variable over the course of their infection. Also, different individuals shed different amounts of virus depending, potentially, on the severity of their infection, their vaccination status, and whether they’ve had a prior infection (Zeiss, Asher, Vander Wyk, Allore, & Compton, 2021).

Shedding of virus in wastewater by infected vaccinated individuals was observed in our study. On several occasions, the entire unvaccinated population of a barracks was tested at Training Center B and found to be negative, suggesting that asymptomatic vaccinated individuals were responsible. On one occasion, the entire population was vaccinated. When individuals were not clinically diagnosed following a rise in wastewater levels of SARS-CoV-2, elevated levels were detected for a week or longer consistent with continued shedding by an infected individual (Figures 2 and 3).

Testing of wastewater from cutters was easily accomplished by dilution of the blackwater and use of the lipophilic iron to capture viral particles as described in the Methods. However, testing blackwater faces many logistical challenges. While at sea, storage of the samples and their delivery to testing laboratories is one challenge. Adapting blackwater systems to allow for sampling is another hurdle, as hardware modification is required to facilitate sampling. While in port, individuals often do not live on the vessels and may acquire SARS-CoV-2 at other locations. Significant alterations to the normal operational flow of cutters would be required to make wastewater testing an alternative to regular screening of individuals by antigen or PCR tests.

Overall, wastewater testing is a useful epidemiological tool. It is non-invasive and much less resource-intensive relative to mass screening and may be of particular use when monitoring a vaccinated population that is not regularly tested by other means. Our work shows several instances of detection of infections in asymptomatic individuals which resulted in isolation that prevented subsequent spread of the disease, highlighting the value of this important tool in combatting SARS-CoV-2 and other infectious diseases (Lappan et al., 2021; Zahedi, Monis, Deere, & Ryan, 2021).

## Data Availability

All data produced in the present study are available upon reasonable request to the authors. However, locations of military training centers will not be identified.

## Acknowledgements

The authors thank the many collaborators with this work and, in particular, the outstanding support of reservist Coast Guard personnel including Erin L. Barttelt, Chad R. Cruset, Erin Degenstein, William Cabreja, Paul Woodhead, Meghan Peterson, Caitlin Springer, Richard Burmeister, Shawn Tuthill, Samuel Encarnacion, Eric Propst, Jeremy Guerrero, Lorenzo Medina, Yevginy Kislov, and Joseph Guajardo. The authors thank Kendra Maas and Lisa Nigro of the Microbial Analysis, Resources, and Services (MARS) Center of the University of Connecticut for their training and support in adopting the testing methodology. The authors thank Michael Raneri for his careful review of the work. The views expressed herein are the opinions of the authors and not necessarily those of the U.S. Coast Guard or federal government.

## Statement of Contributions

J.P.G., G.J.H., E.J.P., and D.L. conceived of and drafted the manuscript. M.R., C.H., A.K. E.L., A.R., J.P.G., N.P., S.Z., C.F., S.G., D.L., and A.D. designed sampling methodology, obtained samples and/or performed laboratory work. G.J.H., E.J.P., D.T., B.A.O., J.I., I.K., E.B., and J.P. provided materiel and leadership support.

